# Epidemiological characteristics of mpox virus Clade Ib in the Democratic Republic of the Congo: the impact of transmission mode

**DOI:** 10.1101/2025.05.27.25328406

**Authors:** Cécile Kremer, Sabin Sabiti Nundu, Emmanuel Hasivirwe Vakaniaki, Isabel Brosius, Guy Mukari, Papy Munganga, Eugene Bangwen, Jean Claude Tshomba, Yves Mujula, Elise De Vos, Christophe Van Dijck, Sarah Houben, Oswaldo Gressani, Cris Kacita, Daniel Mukadi-Bamuleka, Tony Wawina, Eddy Lusamaki, Amuri Aziza, Patrick Mutombo, Jean-Jacques Muyembe-Tamfum, Placide Mbala-Kingebeni, Laurens Liesenborghs, Niel Hens, Andrea Torneri

## Abstract

Monkeypox virus (MPXV) Clade Ib, a new MPXV subclade associated with sustained human-to-human transmission, emerged in eastern DRC in late 2023. Early spread mainly occurred among the adult population, primarily through sexual contact. We investigated how key epidemiological characteristics of the new MPXV Clade Ib outbreak in the DRC differ by transmission mode. We estimated the mean serial interval to be 8.57 days (95% CI 7.15 – 9.91 days). The serial interval was approximately four days shorter for putative sexual compared to non-sexual transmission. We estimated the mean incubation period to be 9.56 days (95% CrI 8.22 – 11.02 days), and approximately two days shorter for putative sexual compared to non-sexual transmission. These observed differences in serial interval and incubation period estimates by transmission mode suggest that the context in which an outbreak occurs is an essential factor to consider when devising public health policies.

## Introduction

Mpox is a zoonotic viral disease characterized by fever, lymphadenopathy, and a vesiculopustular rash, caused by the mpox virus (MPXV, formerly Monkeypox virus), an orthopoxvirus related to variola virus. In late 2023, after decades of increasing zoonotic outbreaks of MPXV in the Democratic Republic of the Congo (DRC), a new subclade of MPXV Clade I associated with sustained human-to-human transmission emerged in eastern DRC. This subclade first appeared in Kamituga, a mining town in South Kivu province, and spread primarily - but not exclusively - through heterosexual contact [1]. Although most infections occurred among adults, one fifth of patients were children who got infected within the household [1, 2]. Later reports from surrounding areas found that non-sexual contact was the predominant mode of transmission [3]. From South-Kivu, MPXV Clade Ib spread to other regions of the DRC, and to neighboring countries, prompting the WHO to declare MPXV a public health emergency of international concern, in August 2024.

Effective epidemic response strategies require an understanding of transmission dynamics and important epidemiological delays, including the serial interval (i.e., the time between symptom onset in a primary case and symptom onset in a secondary case) and incubation period (i.e., the time between infection and symptom onset). These delays are directly linked to epidemic growth. A shorter serial interval allows for more rapid transmission between individuals, and when combined with a short incubation period this can lead to faster epidemic expansion. These delays influence the basic reproduction number and the doubling time of MPXV, making them key determinants of how quickly an epidemic grows and how intensively interventions must be applied [4]. Since these delays are not fixed for a given pathogen but may differ by genomic lineage, context, or mode of transmission, investigating how they differ by transmission mode is important as it will affect the effectiveness of control measures.

These key epidemiological delays have been estimated for MPXV Clade IIb, another subclade of MPXV which emerged in Nigeria and started a global outbreak in 2022 [5]. This ongoing outbreak primarily affects men who have sex with men, and the primary mode of transmission is sexual contact. The estimated mean serial interval for sexually transmitted MPXV Clade IIb was 10.1 days (95% CrI 6.6 - 14.7 days) in the Netherlands [6]. The estimated incubation period was on average 9.1 days (95% CI 6.5 - 10.9 days) and the estimated mean generation time was 12.5 days (95% CI 7.5 - 17.3 days) among MPXV Clade IIb patients in Italy [7]. One study in the UK found that the median serial interval was shorter than the incubation period, indicating considerable presymptomatic transmission of MPXV Clade IIb [8], a finding that was later corroborated by virological shedding data [9].

Shorter incubation periods have been observed after exposure to broken skin or mucous membranes during the first outbreak of human mpox outside of Africa [10]. Likewise, studies have also described varying serial intervals for MPXV by context and mode of transmission. Based on pooled estimates, the serial interval for MPXV Clade IIb was shorter during the 2022 outbreak in Europe compared to historical outbreaks of MPXV Clade Ia. This shortening of the serial interval possibly corresponds to differences in the mode of transmission and contact patterns [11]. A study on MPXV Clade Ia using data from several endemic settings reported considerably longer mean serial intervals in case of household transmission compared to hospital transmission, suggesting variability in serial intervals depending on the outbreak setting [12].

Whether similar estimates apply to outbreaks of MPXV clade Ib has not yet been extensively studied. In one study of Clade Ib in Uvira, DRC, the incubation period was shorter after sexual versus non-sexual transmission. In addition, incubation periods were shorter in the beginning of the outbreak compared to the later phase - a shift that was paralleled by an increase in non-sexually transmitted infections over time [13]. To our knowledge, there are no studies that have estimated the serial interval of MPXV Clade Ib by transmission route. Here, we aimed to estimate the serial interval and incubation period distributions for MPXV Clade Ib and to determine how they differ by transmission mode.

## Results

### Participant characteristics

We conducted a secondary analysis of data from a prospective clinical characterization study of individuals infected with MPXV Clade Ib (see Methods) [2]. Between May 2024 and February 2025, 1013 participants (650 from Kamituga and 363 from Goma) tested positive on MPXV-PCR and were included in the current study. Of these, 503 (49.66%) were female, 578 (57.06%) were adults (≥ 18 years), 79 (7.80%) were aged 12-17 years, and 356 (35.14%) were below 12 years of age. The median age was 20 years (IQR 5 - 26 years; Supplementary Figure S1). Among adults, the most commonly reported occupations were mine worker (122/578, 21.11%), trader (98/578, 16.96%), sex worker (74/578, 12.80%), law enforcement (43/578, 7.44%), and farmer (40/1013, 6.92%). The majority (795/1013, 78.48%) of participants reported having at least one other household member with mpox. Cutaneous lesions were present on the genitals in 329 (67.14%) women and in 332 (66.67%) men. Of the 657 MPXV-infected individuals older than 12 years, 475 (72.30%) reported sexual intercourse in the preceding three weeks. Among these, 162 (34.11%) reported sexual contact with a sex worker. The data from the Kamituga cohort have been described in detail previously [2].

### Key transmission parameters

To estimate the serial interval and incubation period distributions, we used data on self-reported contact between study participants and other individuals infected with MPXV collected during the baseline visit in the MBOTE study (see Methods). Of the 650 participants in the Kamituga cohort, 344 (52.92%) reported to have had contact with a confirmed or suspected mpox patient. In 157/344 (45.64%), the reported contact was a participant of the same study, allowing us to pair study participants. Among these 157 individuals, 111 (70.70%) reported only one contacted individual, of which 48 (43.24%) were sexual contacts. Of the 313 participants in the Goma cohort, 71 (22.68%) reported to have had contact with a confirmed or suspected mpox patient, of which 16/71 (22.54%) were a participant of the same study. Due to the limited available transmission pairs in Goma, these individuals were not included in the serial interval estimation. Of the 1000 MPXV-infected individuals from Kamituga and Goma for whom the date of symptom onset was available, 158 reported contact with at least one other mpox patient as well as the date of last contact with that patient. Of those, 114 (72.15%) reported only one contact, of which 20 (17.54%) were a single exposure.

#### Serial interval

We used the R package EpiDelays [14] to obtain a non-parametric estimate of the serial interval distribution among the 111 transmission pairs from Kamituga where the MPXV-infected individual reported contact with only one other study participant. Overall, the mean serial interval was estimated to be 8.57 days (95% CI 7.15 - 9.91 days; Table 1). To account for missing data on the presumed mode of transmission (i.e., sexual versus non-sexual) we included three scenarios, where the missing transmission mode was assumed to be either sexual or non-sexual (see Methods). The estimated serial interval was consistently shorter for sexual transmission in all three scenarios (e.g. 6.86 days [95% CI 5.25 – 8.68 days] vs. 9.97 days [95% CI 7.82 – 12.13 days]; Table 1). In addition, the serial interval for non-sexual transmission was shorter when the secondary case was a child under 12 years of age compared to adult secondary cases (8.27 days [95% CI 5.45 – 11.41 days] vs. 10.82 days [95% CI 8.22 – 13.50 days]; Table 1). When including only participants older than 12 years, the serial interval was approximately 4 days shorter for sexual transmission (6.86 days [95% CI 5.14 – 8.59 days] vs. 10.82 days [95% CI 8.22 – 13.50] days; Table 1). Presymptomatic transmission was observed in 7 pairs (6.31% of all transmission pairs), of which 4 (57.14%) reported sexual contact. Among 48 reported sexual contacts, 27 (56.25%) were with a household member, 12 (25.00%) were related to professional sex work, 5 (10.42%) were with a friend, and 4 (8.33%) with an unspecified sexual partner. Of the 40 reported non-sexual contacts, 18 (45.00%) were with a household member, 4 (10.00%) with another family member, 17 (42.50%) with a colleague, and 1 (2.50%) with a neighbor. Of the 23 transmission pairs for which information on the sexual nature of the contact was not available, 20 (86.96%) were with a household member, 1 (4.35%) with another family member, 1 (4.35%) with a colleague, and 1 (4.35%) with a neighbor. Of these, 10 (43.48%) individuals did present with cutaneous genital and/or anal lesions at the baseline visit.

**Table 1.** Non-parametric estimates of the serial interval mean and standard deviation (SD).

|  | Mean (95% CI) | SD (95% CI) | Mean (95% CI) | SD (95% CI) |
| --- | --- | --- | --- | --- |
| **All pairs** | 8.57 (7.15 – 9.91) | 7.46 (6.45 – 8.35) |  |  |
|  | *Sexual transmission* |  | *Non-sexual transmission* |  |
| **Scenario 1** | 6.86 (5.25 – 8.68) | 6.12 (4.67 – 7.50) | 9.97 (7.82 – 12.13) | 8.13 (6.72 – 9.20) |
| **Scenario 2** | 6.96 (5.31 – 8.77) | 6.20 (4.69 – 7.47) | 9.79 (7.94 – 11.76) | 8.07 (6.71 – 9.09) |
| **Scenario 3** | 6.86 (5.12 – 8.61) | 6.18 (4.73 – 7.51) | 9.92 (7.97 – 11.95) | 8.07 (6.74 – 9.16) |
|  | *Child (<12y) secondary case* |  | *Adult/adolescent secondary case* |  |
| **Non-sexual** | 8.27 (5.45 – 11.41) | 7.21 (4.02 – 9.39) | 10.82 (8.22 – 13.50) | 8.38 (6.83 – 9.56) |
|  | *Sexual transmission (≥ 12y)* |  | *Non-sexual transmission (≥ 12y)* |  |
| **Scenario 1** | 6.86 (5.14 – 8.59) | 6.16 (4.71 – 7.52) | 10.82 (8.22 – 13.50) | 8.38 (6.83 – 9.59) |
| **Scenario 2** | 6.96 (5.31 – 8.77) | 6.20 (4.69 – 7.47) | 10.61 (8.12 – 13.27) | 8.39 (7.00 – 9.58) |
| **Scenario 3** | 6.86 (5.14 – 8.59) | 6.18 (4.71 – 7.52) | 10.82 (8.22 – 13.50) | 8.38 (6.83 – 9.56) |

We used a Bayesian linear regression model to investigate the influence of sexual and household transmission on the mean serial interval while accounting for age (see Methods). Of all participants with known symptom onset, 157 MPXV-infected individuals reported contact with at least one other patient included in the study, resulting in 260 individuals (i.e. primary and secondary cases) being included in the serial interval estimation. We found no evidence for a difference in serial intervals for household vs. non-household transmission or for children vs. adults (Table 2). The serial interval was 3.72 days (95% CrI 6.62 to 0.71 days) shorter for sexual vs. non-sexual transmission (Table 2). Of the 126 source cases in the most likely transmission network (Figure 1), 23 (18.25%) were female sex workers and 25 (19.84%) were mine workers. The majority of known source cases were women (54.76%) and transmission occurred via sexual contact in 39.58% of infector-infectee pairs, of which 56.14% were household members.

**Figure 1.**
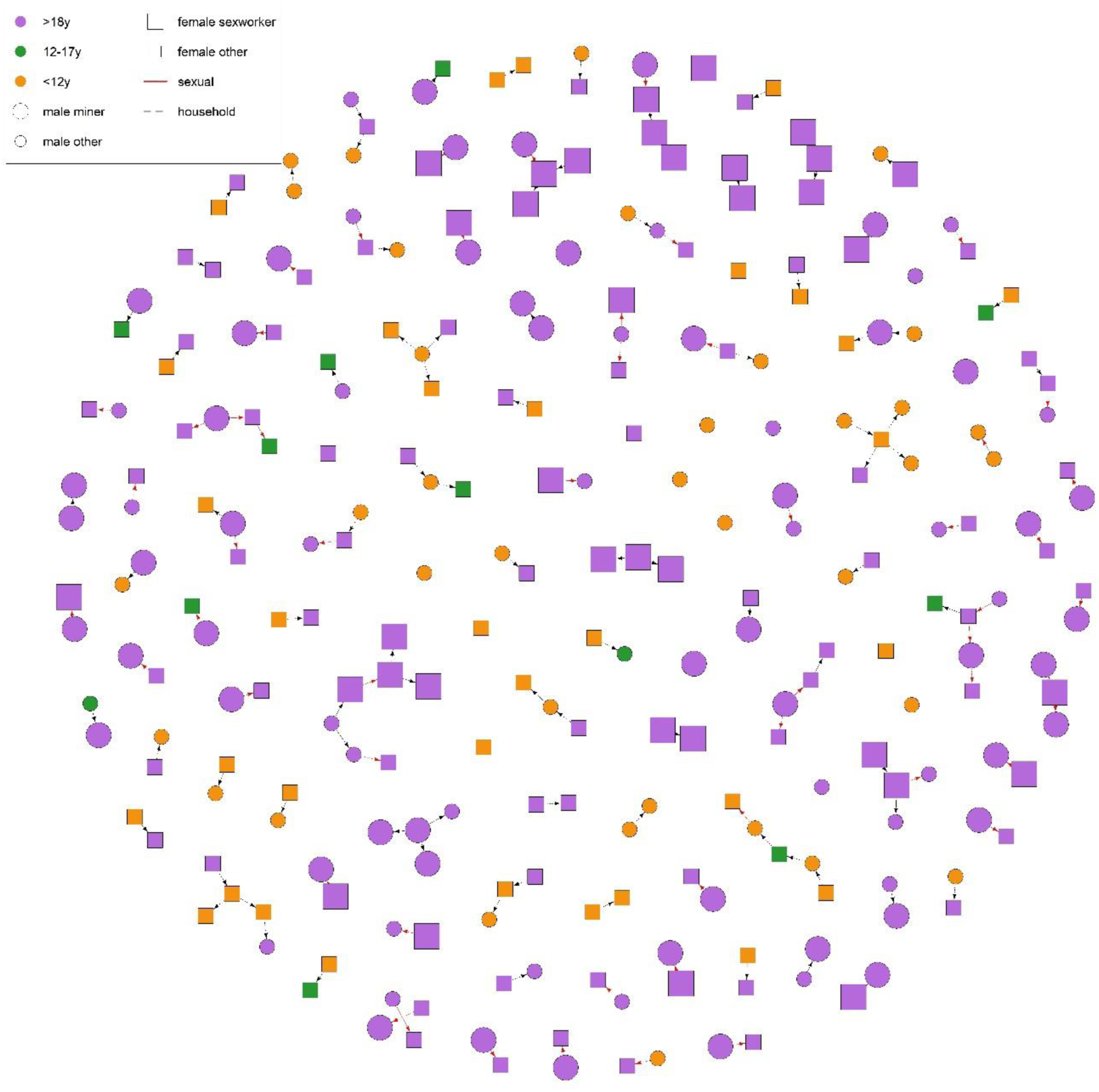
Most likely transmission network based on the Bayesian linear regression model.

**Table 2.** Regression coefficients of the Bayesian linear model for the normally distributed serial interval.

| Parameter | Median | 95% CrI |
| --- | --- | --- |
| Intercept | 9.95 | 6.31 to 13.56 |
| Sexual transmission ($\beta_1$) | -3.72 | -6.64 to -0.71 |
| Adult (>12y) secondary case ($\beta_2$) | 1.48 | -2.06 to 5.07 |
| Household transmission ($\beta_3$) | -1.56 | -4.40 to 1.33 |
| Standard deviation $\sigma$ | 7.70 | 6.85 to 8.74 |

#### Incubation period

We estimated the incubation period of MPXV Clade Ib following the Bayesian approach used by Ward et al. [8], assuming a Weibull distribution (see Methods). We used the same scenarios for sexual transmission as for the observed serial intervals. Among the 114 MPXV-infected individuals who reported contact with a single mpox patient, the mean incubation period was estimated to be 9.56 days (95% CrI 8.22 - 11.02 days). The incubation period was consistently shorter with about 2 days for putative sexual transmission compared to non-sexual transmission (e.g. 8.51 days [95% CrI 6.84 – 10.54 days] vs. 10.81 days [95% CrI 8.75 – 12.96 days], Table 3). Sensitivity analyses increasing (decreasing) the lower bound of 21 days resulted in slightly higher (lower) estimates of the mean incubation period (Supplementary Table S1). On average, onset of rash occurred 1.47 days (median 1 day, IQR 0 - 2 days) after reported symptom onset.

**Table 3.** Estimated mean and standard deviation (SD) of the incubation period, based on a Weibull distribution accounting for censoring and right truncation.

|  | Mean (95% CrI) | SD (95% CrI) | Mean (95% CrI) | SD (95% CrI) |
| --- | --- | --- | --- | --- |
| **Overall** | 9.56 (8.22 – 11.02) | 5.55 (4.60 – 6.84) |  |  |
|  | *Sexual transmission* |  | *Non-sexual transmission* |  |
| **Scenario 1** | 8.51 (6.84 – 10.54) | 5.06 (3.79 – 7.02) | 10.81 (8.75 – 12.96) | 5.65 (4.41 – 7.65) |
| **Scenario 2** | 8.59 (6.75 – 10.91) | 5.19 (3.79 – 7.51) | 10.42 (8.57 – 12.41) | 5.57 (4.45 – 7.34) |
| **Scenario 3** | 8.71 (6.92 – 10.88) | 5.10 (3.75 – 7.21) | 10.38 (8.48 – 12.42) | 5.72 (4.53 – 7.69) |
|  | *Child (< 18y) secondary case* |  | *Adult ($\geq 18y$) secondary case* |  |
| **Non-sexual** | 11.44 (8.92 – 14.07) | 4.49 (3.25 – 6.93) | 10.15 (7.36 – 13.88) | 7.03 (4.87 – 12.59) |
|  | *Child (<12y) secondary case* |  | *Adult/adolescent secondary case* |  |
| **Non-sexual** | 11.11 (7.89 – 14.36) | 4.71 (3.42 – 6.93) | 9.08 (6.37 – 11.90) | 6.24 (4.54 – 9.71) |
|  | *Sexual transmission* |  | *Non-sexual transmission* |  |
| **Adults only** | 8.43 (6.56 – 10.80) | 5.24 (3.79 – 7.71) | 10.15 (7.33 – 13.94) | 7.01 (4.80 – 12.72) |
| **Adults + adolescents** | 8.59 (6.73 – 10.88) | 5.19 (3.79 – 7.52) | 10.16 (7.75 – 13.03) | 6.26 (4.56 – 9.72) |

We used a Bayesian linear regression model to investigate the influence of sexual transmission and age on the incubation period (see Methods). We found no substantial evidence for a difference in incubation periods for putative sexual versus non-sexual transmission, age group, or their interaction (all 95% CrI include 0; Table 4). Compared to Goma, the evidence for a shorter incubation period for sexual transmission was stronger in Kamituga (lower *γ_1_* and 95% CrI closer to 0; Table 4).

**Table 4.** Regression coefficients (median [95% CrI]) of the Bayesian linear model for the Weibull distributed incubation period (N = sample size).

|  | Kamituga (N = 81) | Goma (N = 33) | Combined (N = 114) |
| --- | --- | --- | --- |
| Intercept | 10.86 (7.61 to 14.91) | 9.84 (5.47 to 17.20) | 10.70 (8.04 to 13.88) |
| Sexual transmission ($\gamma_1$) | -2.15 (-5.25 to 0.90) | -0.75 (-8.21 to 8.66) | -1.83 (-4.26 to 0.96) |
| Age $\geq 12y$ ($\gamma_2$) | 0.60 (-3.40 to 3.84) | -0.94 (-8.55 to 7.61) | -0.24 (-3.60 to 2.76) |
| Weibull shape $\alpha$ | 2.16 (1.69 to 2.75) | 1.06 (0.59 to 1.68) | 1.81 (1.46 to 2.22) |

OPX-specific cycle threshold (Ct) values obtained from skin lesion samples were available for 960 MPXV-infected individuals from Kamituga and Goma (Supplementary Figure S2). Ct values were available for 110 secondary cases included in the serial interval estimation. Using a linear regression model that accounts for the time between rash onset and sample date, we found that Ct values were significantly lower for individuals reporting sexual contact as the presumed transmission mode (Table 5). Among secondary cases included in the incubation period estimation, Ct values were available for 6 individuals from Goma and 50 individuals from Kamituga. Among those, older individuals had significantly lower Ct values and Ct values were lower in Kamituga (Table 5).

**Table 5.** Regression coefficients of the linear model for Ct values, including transmission pairs used in the serial interval estimation (N = 110) or secondary cases used in the incubation period estimation (N = 56).

|  | Transmission pairs (serial interval) |  | Secondary cases (incubation period) |  |
| --- | --- | --- | --- | --- |
|  | Estimate (SE) | *p*-value | Estimate (SE) | *p*-value |
| Intercept | 26.89 (1.96) |  | 34.77 (3.88) |  |
| Days between rash onset and sample | -0.06 (0.29) | .840 | -0.003 (0.32) | .994 |
| Sexual transmission | -4.51 (1.47) | .003 | -2.75 (2.16) | .209 |
| Age | -0.08 (0.07) | .236 | -0.25 (0.10) | .017 |
| Kamituga site | n.a. | n.a. | -6.48 (3.22) | .049 |

## Discussion

We estimated the overall mean serial interval of MPXV Clade Ib to be 8.57 days (95% CI 7.15 – 9.91 days), and we found that the mean serial interval for putative sexual transmission of MPXV Clade Ib was approximately three days shorter than for non-sexual transmission. When excluding children under 12 years of age from the analysis, for whom transmission was always presumed to be non-sexual, the difference in serial intervals between sexual and non-sexual transmission was as large as four days. The shorter serial interval estimated for putative sexual transmission indicates that MPXV Clade Ib is transmitted more rapidly through sexual contact. This could be because during sexual contact, large quantities of virus particles shed from genital lesions come into direct contact with the genital, oral, or anorectal mucosa, facilitating rapid infection. Consistent with this hypothesis of higher viral shedding, we found that Ct values among individuals in known transmission pairs were significantly lower for those who reported sexual contact as the presumed transmission route, indicating higher viral loads. Other important factors contributing to observed differences in serial intervals are the contact process and the risk of infection for a given type of interaction. Future work should consider the use of simulation models to investigate this.

Our overall estimate of the mean incubation period (9.56 days [95% CrI 8.22 – 11.02 days]) is substantially lower than what was recently found among MPXV Clade Ib patients in Uvira, DRC [13]. However, we found a mean incubation period of 8.51 days (95% CrI 6.84 – 10.54 days) for sexual transmission, which is similar to the previous estimate from Uvira [13]. In our study, the mean incubation period was approximately two days shorter for putative sexual compared to non-sexual transmission. We found that the incubation period was slightly longer for children under 12 years of age, implying that age of the secondary case could be an important confounder when estimating the serial interval and incubation period for sexual and non-sexual MPXV Clade Ib transmission, which has not been accounted for in the previous incubation period estimate from Uvira [13]. Evidence for a shorter incubation period for putative sexual transmission was stronger among patients from Kamituga compared to Goma, which could imply that sexual transmission was more important in Kamituga due to the higher presence of professional sex work and consequently sex workers probably played a significant role in transmission in Kamituga.

This study has several limitations. First, the RADI platform was used for initial screening of skin lesion samples. Clade-specific PCR or sequencing was not performed, which limits our ability to confirm with certainty that all detected cases belong to Clade Ib. However, the epidemiological context strongly suggests Clade Ib circulation in both study sites. In addition, in a subset of 253 participants, we previously confirmed that they were infected with Clade Ib using a lad-specific PCR [2]. Second, the distinction between sexual and non-sexual transmission is based on self-reporting of sexual contact. The high prevalence of genital and/or anal lesions in our study population suggests that sexual transmission may have been more common than reported. Third, we did not have information on the precise location of skin lesion samples, and specific Ct values for genital samples are pending. Variations in Ct values between locations could exist, especially between genital and non-genital lesions. Fourth, for estimation of the incubation period only the last date of contact was known. Therefore we had to resort to using relatively wide exposure windows for individuals who reported multiple exposures to the same mpox patient, while the method assumes that the probability of infection during this interval is approximately constant [13, 15]. We found that our estimates are slightly sensitive to different exposure window widths. However, the comparison of sexual versus non-sexual transmission is expected to be robust. Future work could benefit from utilizing transmission dynamics to allow for a probabilistic exposure window, e.g. based on longitudinally collected viral load data.

Differences in key epidemiological characteristics such as the serial interval and incubation period between putative sexual and non-sexual transmission are highly relevant for public health policy. Shorter serial intervals indicate faster transmission, hence case detection and intervention measures such as targeted vaccination should be implemented more rapidly in areas where MPXV Clade Ib transmission is driven by sexual contact. These differences also suggest that the transmission mode must be considered when devising policies regarding isolation periods and quarantine recommendations. Accurate estimates of these epidemiological characteristics also improve transmission models, enabling better prediction of outbreak dynamics and the impact of targeted public health strategies. Although MPXV Clade Ib was first thought to be transmitted mainly through sexual contact, the shift in age of affected individuals back to younger individuals in other areas indicates that the context of the outbreak setting is an essential factor to consider when devising public health policy [16]. In light of this, there is a need for systematic data collection in different areas in order to characterize the changing epidemiology of MPXV.

## Methods

### Study design and data collection

The main objective of the present study was to obtain estimates of the serial interval and incubation period distributions for MPXV Clade Ib. To this end, we conducted a secondary analysis of data from a prospective clinical characterization study of individuals infected with MPXV Clade Ib: the Mpox Biology, Outcome, Transmission, and Epidemiology – Kamituga study (MBOTE-Kamituga; ClinicalTrials.gov: NCT06652646; www.mbote-mpox.com). This study started in May 2024 and took place in Kamituga Health Zone, South-Kivu province, the initial epicenter of the MPXV Clade Ib outbreak. The study setup and initial findings have been published previously [2]. In brief, individuals presenting with suspected MPXV Clade Ib infection were invited to participate. After providing written informed consent, participants completed a baseline survey prior to diagnostic testing. This survey captured demographic information, medical history, clinical symptoms, date of symptom onset, and potential exposure to animals. In addition, exposure to other MPXV-infected individuals was assessed in depth, including the timing and nature of the contact. Questions about sexual behavior were restricted to individuals over 12 years old. If a participant reported contact with another MPXV-infected individual who was also included in the study, this was recorded so that infector-infectee pairs could be identified.

Diagnostic testing was conducted on site using Xpert® Mpox cartridges on a four-module GeneXpert real-time PCR system (Cepheid, Sunnyvale, CA, USA). Participants who tested positive for MPXV by PCR were followed longitudinally throughout their hospital stay. Following discharge, they were invited to attend two outpatient follow-up visits at 29 and 59 days after confirmation of infection.

As the outbreak progressed beyond South Kivu to other provinces, affecting the city of Goma, additional sites were added in Goma from November 2024 onwards. Participant recruitment was conducted using a case report form consistent with that of Kamituga, with context-specific adaptations to Goma. Samples were transported from study sites to the INRB referral laboratory in Goma where MPXV testing was conducted using the RADI PCR platform (KH Medical, Seoul, South Korea). Unlike Kamituga, no longitudinal follow up of confirmed cases was conducted in Goma.

### Estimating key transmission parameters

#### Serial interval

We used the R package EpiDelays (https://github.com/oswaldogressani/EpiDelays) [14] to obtain a non-parametric estimate of the serial interval distribution among transmission pairs where the MPXV-infected individual identified only one contact in the study. The serial interval was constrained to be between -5 and 30 days [8]. Confidence intervals were computed based on 2000 bootstrap replicates of the data. We also estimated the serial interval distribution for sexual and non-sexual transmission under three scenarios: 1) when information on whether the contact was sexual was missing, this was set as sexual contact if the relationship was not direct family (except for spouses), 2) when information on whether the contact was sexual was missing, this was set as a non-sexual contact, and 3) when information on whether the contact was sexual was missing, this was set as sexual contact whenever genital and/or anal cutaneous lesions were present in the secondary case and the relationship was not direct family (except for spouses).

In addition, we used a Bayesian linear regression model to investigate the influence of sexual and household transmission on the mean serial interval, while accounting for age. The serial interval *y_j_* was assumed to follow a Normal distribution with mean *β^T^**X*** and standard deviation *σ*. Here, *β^T^**X*** is the linear predictor with ***X*** including sexual transmission (*X_1_*), age group (*X_2_*), household status (*X_3_*), and an interaction between household status and sexual transmission (*X_4_*).

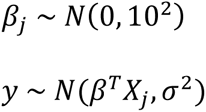

The posterior distribution was evaluated using 5 000 000 iterations with a burn-in period of 40%. Thinning was applied by taking every 200th iteration. Convergence of the MCMC chain was assessed using trace plots (Supplementary Figure S3). The MCMC algorithm consists of two steps that are repeated until convergence: 1) update the transmission tree based on the current parameter values θ^⭑^, and 2) update the parameter values based on the currently accepted transmission tree, using a random-walk Metropolis-Hastings algorithm with a N(θ^⭑^,0.52) proposal distribution. Posterior point estimates of all parameters are given by the 50th percentile of the converged MCMC chain. 95% credible intervals (CrI) are given by the 2.5th and 97.5th percentiles.

For computational reasons, 1000 possible transmission trees were sampled before running the MCMC, and during step 1 of the algorithm one of these trees was randomly selected. Confirmed MPXV patients were linked based on available information on known contacts and specific transmission routes. Transmission trees were sampled as follows:

1. Define possible infectors for each individual based on known contacts. Reported contacts are assumed to be bi-directional (i.e. either individual could have been the index).
2. Impute missing dates of symptom onset by sampling from a uniform distribution with the lower bound set as 21 days before inclusion in the study and the upper bound set as the date of inclusion.
3. Constrain the serial interval to be between -5 and 30 days, consequently updating the list of possible infectors.
4. Assign cluster IDs based on known contacts. Make sure every cluster has an assigned index case to avoid cycles in the network.
5. Sample a transmission tree based on the list of possible infectors (with equal probabilities), taking into account the above constraints.
6. Repeat step 2-6 to obtain 1000 sampled trees.

From all accepted trees, we visualize the most likely transmission tree (i.e. the one that is most often accepted by the MCMC algorithm).

#### Incubation period

We estimated the incubation period of MPXV Clade Ib following the Bayesian approach used by Ward et al. assuming a Weibull distribution [8]. We included all participants with known symptom onset and known date of last contact with an MPXV patient, excluding participants who reported contact with multiple other MPXV patients. For participants that reported a single exposure, the exposure time was fixed to the reported date. To correct for the coarseness of the data (i.e. only the date of exposure is known rather than the exact time), the exposure window for these individuals was defined as the reported date of last contact until the day after. Since most participants reported multiple exposures per contact and only the date of last contact was registered, the time from exposure to symptom onset is right-censored. We therefore set the lower bound of the exposure window for these individuals to 21 days before their symptom onset as this is generally considered as the upper bound of the incubation interval [8], and the upper bound to one day after the reported date of last contact. Participants who reported that the date of last contact occurred after their date of symptom onset were excluded from this analysis. We performed a sensitivity analysis varying the lower bound of the exposure window for individuals reporting repeated exposures.

In addition, we used a Bayesian linear regression model to investigate the influence of sexual transmission and age on the incubation period. The incubation period *y_j_* was assumed to follow a Weibull distribution with mean *γ^T^**X***. Here, *γ^T^**X*** is the linear predictor with ***X*** including sexual transmission (*X_1_*) and age group (*X_2_*), as well as their interaction (*X_3_*).

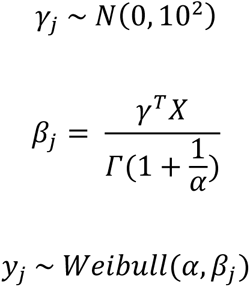

The posterior distribution was evaluated using 4 chains with 10 000 iterations each, with a burn-in period of 50%. Convergence of the MCMC chains was assessed using trace plots.

## Data Availability

Data are available upon request. R code will be made available on GitHub upon publication.

## Code availability

All analyses were performed using R software version 4.3.2. R code is available on GitHub (https://github.com/cecilekremer/mpox_transmission).

## Acknowledgements

AT and LL (project G069725N) and CVD (project 12B1M24N) gratefully acknowledge support from the Research Foundation Flanders (FWO). CK, NH, and OG were supported by the VERDI project (101045989) and OG was supported by the ESCAPE project (101095619), funded by the European Union. Views and opinions expressed are however those of the author(s) only and do not necessarily reflect those of the European Union or the Health and Digital Executive Agency (HADEA). Neither the European Union nor the granting authority can be held responsible for them. The computational resources and services used in this work were provided by the VSC (Flemish Supercomputer Center), funded by the Research Foundation Flanders (FWO) and the Flemish Government department EWI.

## Author contributions

AT, NH, LL, and CK contributed to conceptualization of the study. LL, PM, SSN, EHV, IB, GM, PM, EB, JCT, YM, EDV, CVD, SH, CK, DMB, TW, EL, AA, PM, and JJMT contributed to acquisition of the data. CK, OG, and AT contributed to the analysis code. CK performed the analysis. CK, AT, NH, and LL contributed to the interpretation of results. CK and AT drafted the manuscript. All co-authors critically reviewed and revised the manuscript.

## Ethical considerations

The MBOTE-Kamituga study was approved by the Ethics Committee of the University of Kinshasa (approval ID: ESP/CE/78/2024) and the University Hospital Antwerp (ID: 6383). Written informed consent was obtained from all participants aged 18 years and older. For minors, consent was provided by a parent or legal guardian. Adolescents aged 12 to 17 received age-appropriate information and provided written assent.

---

**Supplementary Table S1.**
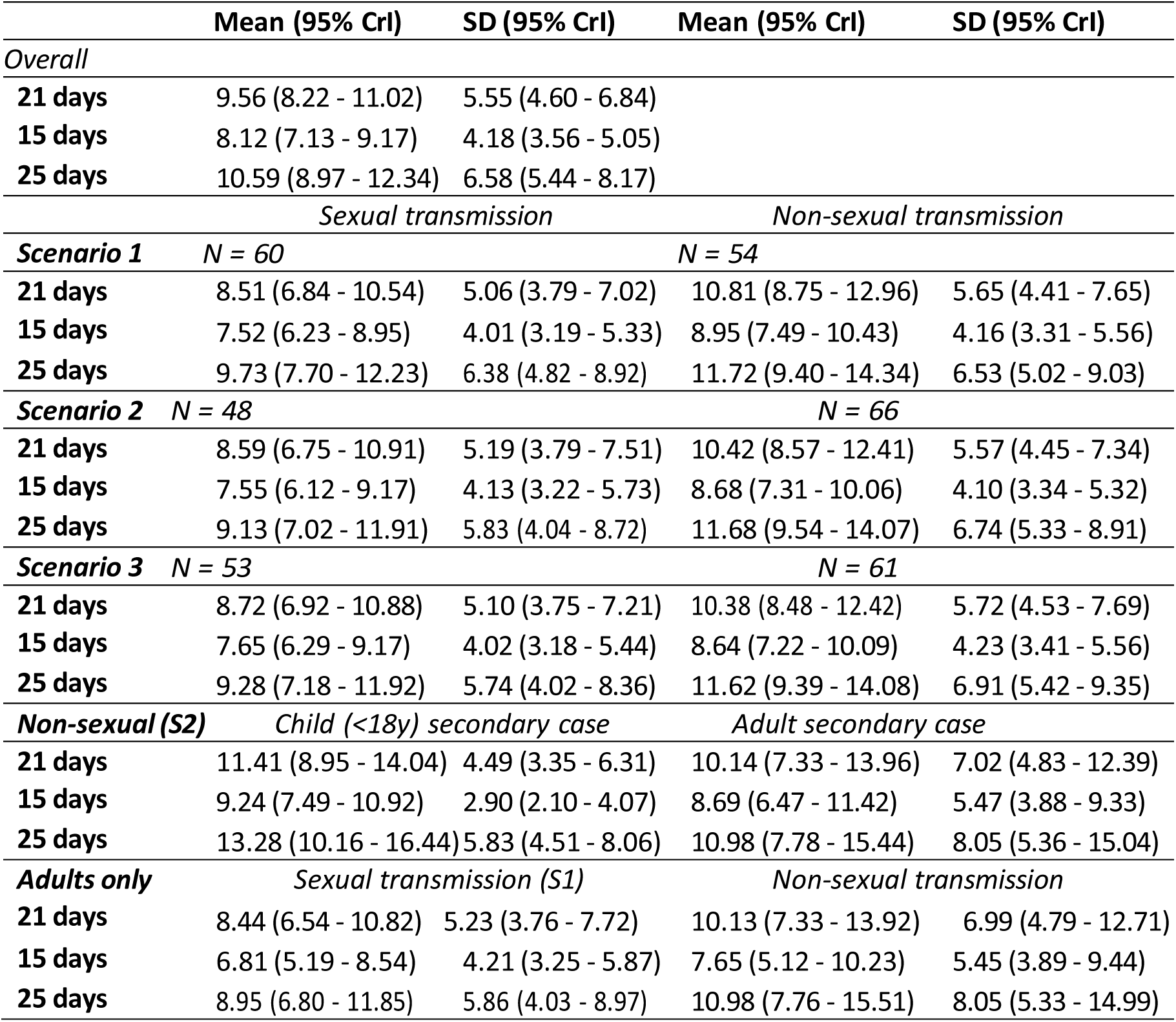
Estimated mean and standard deviation (SD) of the incubation period, based on a Weibull distribution accounting for censoring and right truncation.

|  | Mean (95% CrI) | SD (95% CrI) | Mean (95% CrI) | SD (95% CrI) |
| --- | --- | --- | --- | --- |
| *Overall* |  |  |  |  |
| **21 days** | 9.56 (8.22 - 11.02) | 5.55 (4.60 - 6.84) |  |  |
| **15 days** | 8.12 (7.13 - 9.17) | 4.18 (3.56 - 5.05) |  |  |
| **25 days** | 10.59 (8.97 - 12.34) | 6.58 (5.44 - 8.17) |  |  |
|  | *Sexual transmission* |  | *Non-sexual transmission* |  |
| **Scenario 1** | *N = 60* |  | *N = 54* |  |
| **21 days** | 8.51 (6.84 - 10.54) | 5.06 (3.79 - 7.02) | 10.81 (8.75 - 12.96) | 5.65 (4.41 - 7.65) |
| **15 days** | 7.52 (6.23 - 8.95) | 4.01 (3.19 - 5.33) | 8.95 (7.49 - 10.43) | 4.16 (3.31 - 5.56) |
| **25 days** | 9.73 (7.70 - 12.23) | 6.38 (4.82 - 8.92) | 11.72 (9.40 - 14.34) | 6.53 (5.02 - 9.03) |
| **Scenario 2** | *N = 48* |  | *N = 66* |  |
| **21 days** | 8.59 (6.75 - 10.91) | 5.19 (3.79 - 7.51) | 10.42 (8.57 - 12.41) | 5.57 (4.45 - 7.34) |
| **15 days** | 7.55 (6.12 - 9.17) | 4.13 (3.22 - 5.73) | 8.68 (7.31 - 10.06) | 4.10 (3.34 - 5.32) |
| **25 days** | 9.13 (7.02 - 11.91) | 5.83 (4.04 - 8.72) | 11.68 (9.54 - 14.07) | 6.74 (5.33 - 8.91) |
| **Scenario 3** | *N = 53* |  | *N = 61* |  |
| **21 days** | 8.72 (6.92 - 10.88) | 5.10 (3.75 - 7.21) | 10.38 (8.48 - 12.42) | 5.72 (4.53 - 7.69) |
| **15 days** | 7.65 (6.29 - 9.17) | 4.02 (3.18 - 5.44) | 8.64 (7.22 - 10.09) | 4.23 (3.41 - 5.56) |
| **25 days** | 9.28 (7.18 - 11.92) | 5.74 (4.02 - 8.36) | 11.62 (9.39 - 14.08) | 6.91 (5.42 - 9.35) |
| **Non-sexual (S2)** | *Child (<18y) secondary case* |  | *Adult secondary case* |  |
| **21 days** | 11.41 (8.95 - 14.04) | 4.49 (3.35 - 6.31) | 10.14 (7.33 - 13.96) | 7.02 (4.83 - 12.39) |
| **15 days** | 9.24 (7.49 - 10.92) | 2.90 (2.10 - 4.07) | 8.69 (6.47 - 11.42) | 5.47 (3.88 - 9.33) |
| **25 days** | 13.28 (10.16 - 16.44) | 5.83 (4.51 - 8.06) | 10.98 (7.78 - 15.44) | 8.05 (5.36 - 15.04) |
| **Adults only** | *Sexual transmission (S1)* |  | *Non-sexual transmission* |  |
| **21 days** | 8.44 (6.54 - 10.82) | 5.23 (3.76 - 7.72) | 10.13 (7.33 - 13.92) | 6.99 (4.79 - 12.71) |
| **15 days** | 6.81 (5.19 - 8.54) | 4.21 (3.25 - 5.87) | 7.65 (5.12 - 10.23) | 5.45 (3.89 - 9.44) |
| **25 days** | 8.95 (6.80 - 11.85) | 5.86 (4.03 - 8.97) | 10.98 (7.76 - 15.51) | 8.05 (5.33 - 14.99) |

**Supplementary Figure S1.**
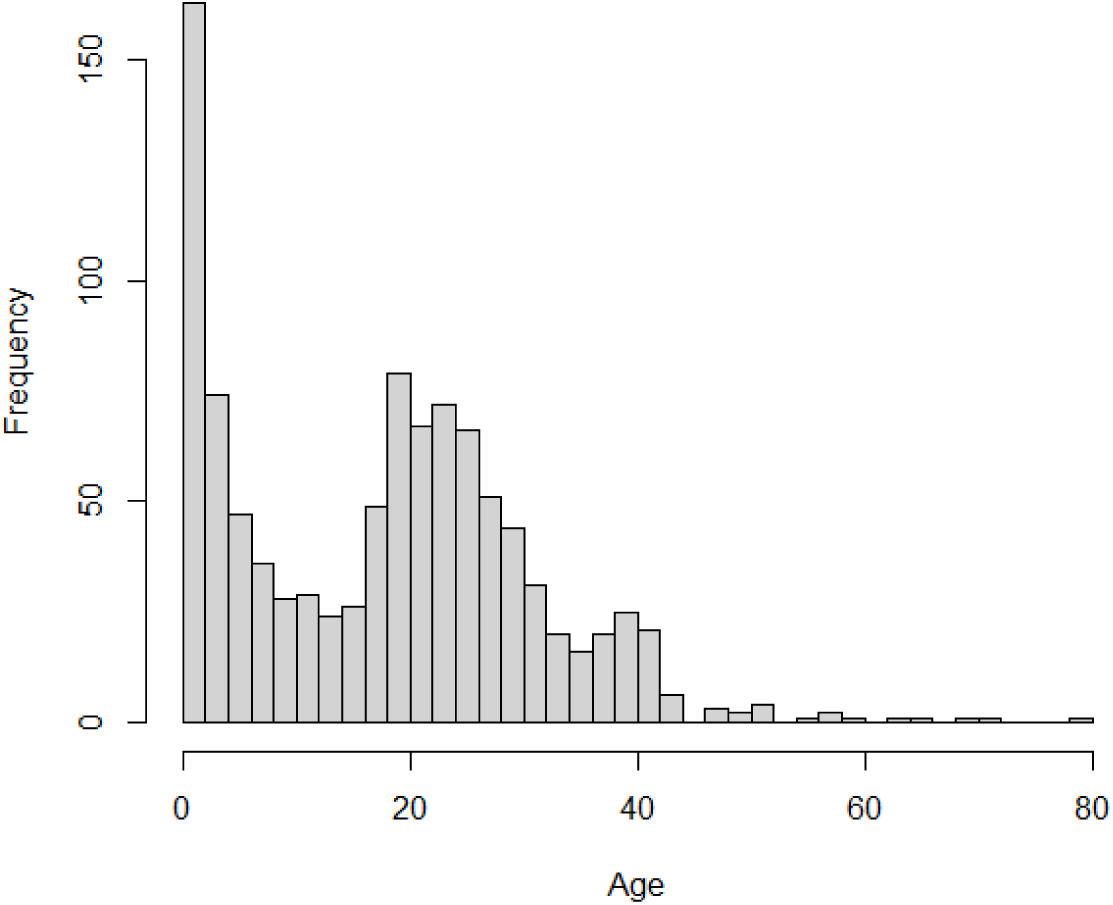
Age distribution of 1013 MPXV-infected individuals from Kamituga and Goma.

**Supplementary Figure S2.**
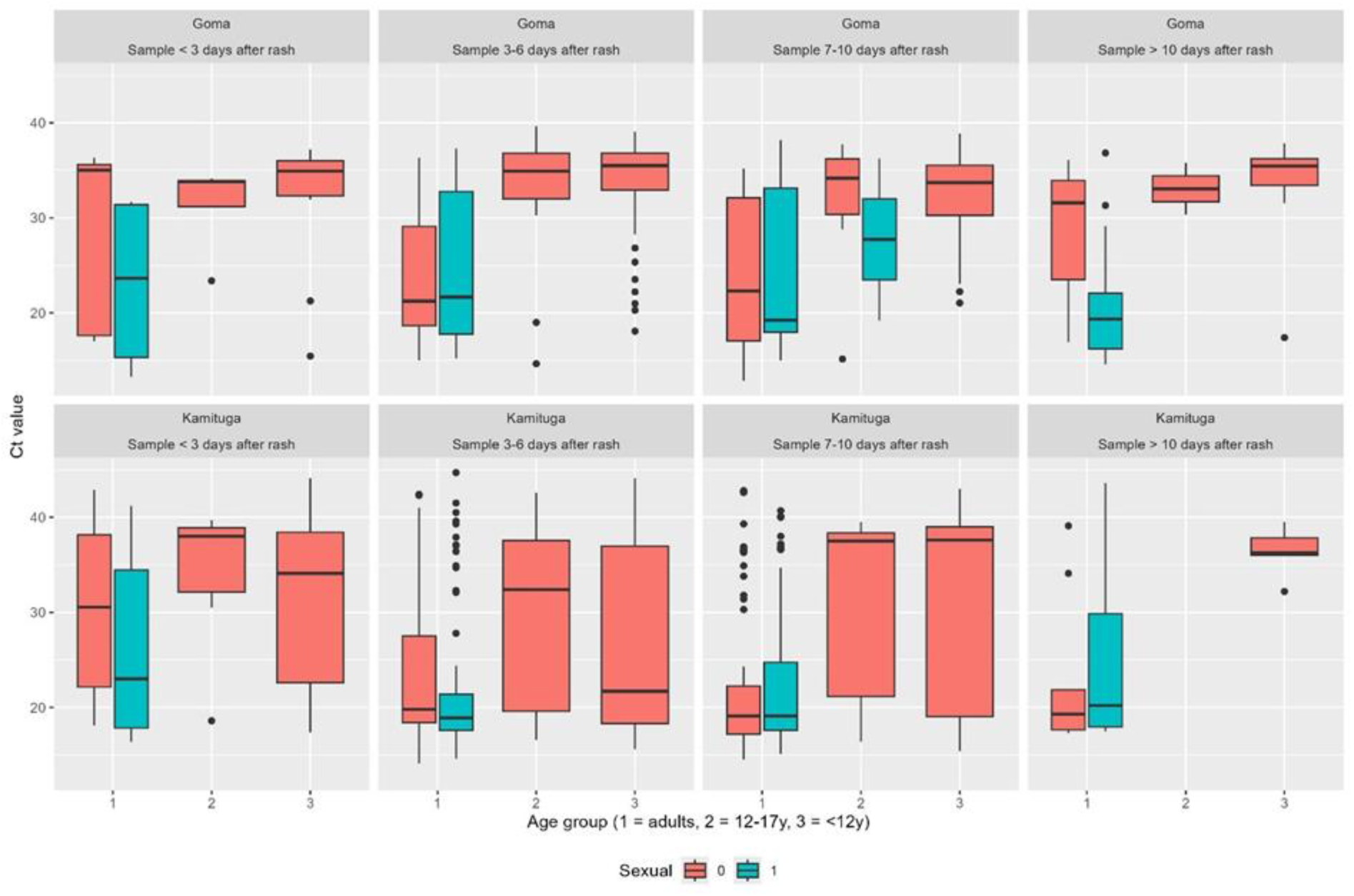
Ct values by location, transmission route, age group, and time between rash onset and sample date.

**Supplementary Figure S3.**
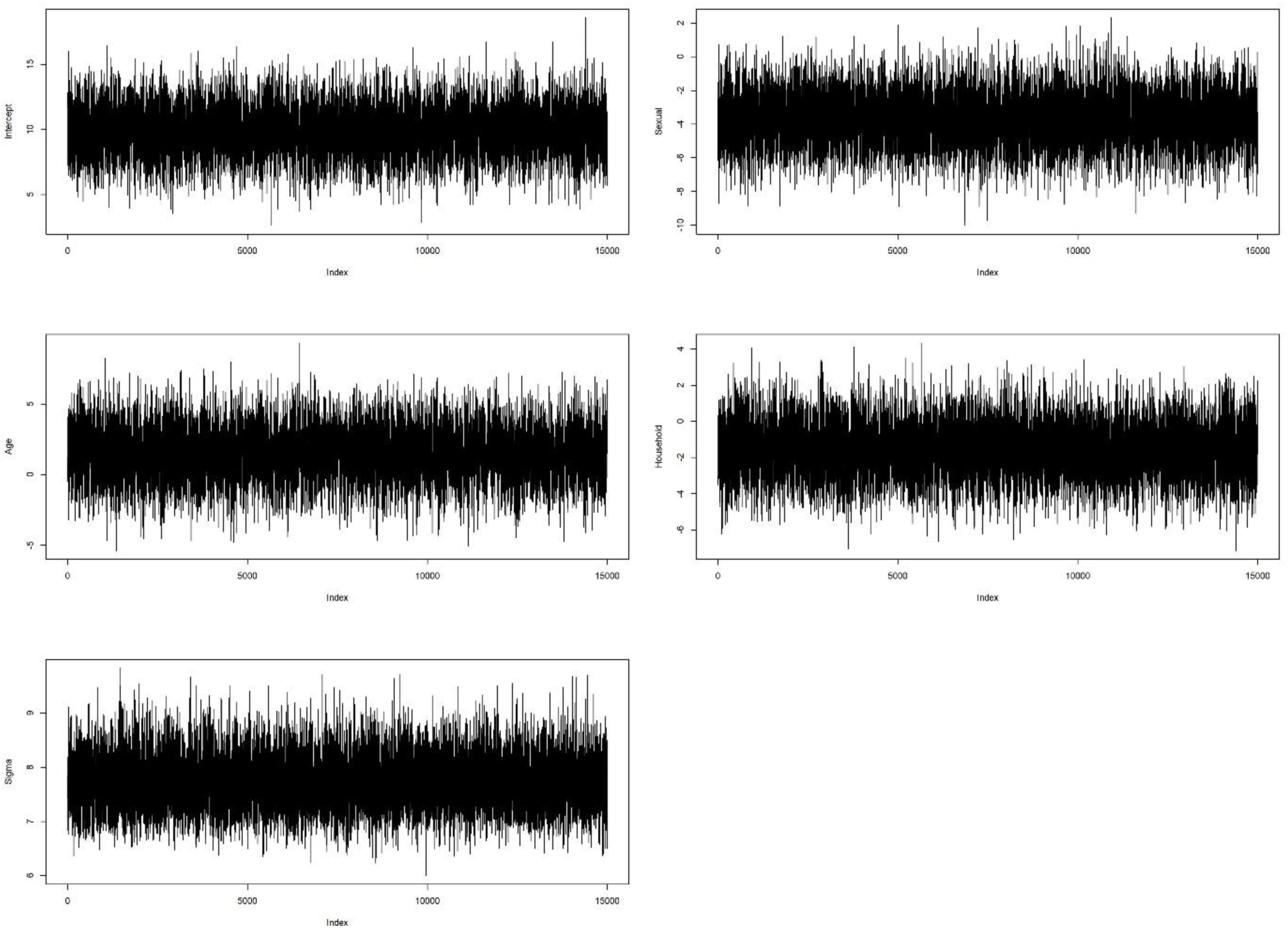
Trace plots of the parameters of the Bayesian regression model for the serial interval.

